# Dietary burden of phosphorus and aluminum in ready-to-eat wheat flour tortillas exceed that of corn tortillas: Implications for patients with renal or cardiovascular disease

**DOI:** 10.1101/2023.09.09.23295298

**Authors:** Kate C. Chiang, Robert A. Yokel, Jason M. Unrine, Kamyar Kalantar-Zadeh, Ajay Gupta

**Author notes:** Corresponding authors Ajay Gupta, M.B.B.S., M.D., Division of Nephrology, Hypertension and Kidney Transplantation, Department of Medicine, University of California Irvine (UCI) School of Medicine, Orange, CA 92868, Robert A. Yokel, Ph.D., Department of Pharmaceutical Sciences University of Kentucky Academic Medical Center, 335 Todd (College of Pharmacy) Building, 789 S. Limestone, Lexington, KY, 40536-0596, US.

## Abstract

Ready-to-eat, shelf-stable tortillas contain several phosphorus- and aluminum-containing additives that may increase the risk of adverse events in patients with chronic kidney disease (CKD). The present study analyzes and compares the elemental content of wheat flour and corn tortillas with special reference to dietary aluminum and phosphorus burden. Twenty-one elements were quantified by ICP-MS and ICP-OES in 14 corn and 13 wheat flour tortilla brands purchased from local supermarkets in Southern California. The aluminum and phosphorus concentrations of many ready-to-eat tortilla brands can present a daily dietary load of up to approximately 100 mg aluminum and 700 mg phosphorus based on an average daily tortilla intake of 330 grams. Ready-to-eat wheat flour tortillas generally had more phosphorus than corn tortillas. Tortillas with aluminum listed as a food additive contained a higher aluminum content than those without such listing, exceeding the tolerable weekly intake. Despite conventional wisdom that CKD patients should avoid phosphorus-rich corn tortillas, ready-to-eat wheat flour tortillas consistently had a higher aluminum and phosphorus content due to additives. CKD patients and healthcare providers should pay attention to food labels, and regulatory authorities should monitor the use of approved food additives and mandate food label warnings for patients at risk.

## Introduction

Phosphorus is the second most abundant mineral in the body and plays a key role in essential biological processes including cell metabolism, energy generation, acid-based balance, and bone calcification.^1^ Grains are the top source of dietary phosphorus in the US diet followed by meat and milk products.^2^ Excess phosphorus intake common to westernized diets^3^ is well-known to be associated with cardiovascular disease and mortality in the general population as demonstrated in the NHANES III study.^4^ With a decrease in urinary phosphorus excretion, renal phosphorus retention is of great concern in patients with chronic kidney disease (CKD).^5^ High serum phosphorus levels and secondary hyperparathyroidism are key contributors to hyperparathyroidism associated bone disease, vascular calcification, and cardiovascular morbidity and mortality in CKD. ^6,7^ Therefore, dietary phosphorus restriction has been recommended for patients with cardiovascular disease or CKD.^8,9^

High prevalence of diabetes and CKD among the Hispanic population have underlined the importance of dietary phosphate restriction in the Latin American diet.^10-12^ The tortilla, originally developed by early Mesoamerican civilizations, is a flat, round, unfermented bread produced from wheat flour or lime (CaO)-cooked maize (corn).^13^ Tortillas are a staple food in Mexico and Central America and have gained widespread popularity around the world, including the US.^13^ In 2001, per capita intake of tortillas was 230-330 grams per day in Mexico, and 16.4 grams per day in the US.^13^ Tortillas are traditionally made fresh at home from flour derived from either corn or maize (Spanish *maíz*) or wheat (Spanish *harina;* Latin *farina*).^14^ However, like other bread products in the last century, tortillas have become commercially available and contain food additives to prolong shelf life and leavening agents to rise and soften the bread.^15,16^ These may provide additional sources of phosphorus and aluminum.^17,18^ Single 6-inch corn and wheat tortillas lacking preservatives contain between 75 to 95 mg and 45 to 65 mg phosphorus, respectively, per 30 grams serving.^19^ Therefore, dietary guidelines for CKD or dialysis patients conventionally recommend wheat tortillas rather than corn tortillas.^19^ However, the phosphorus content of commercially-available corn and wheat tortillas with phosphorus-containing food additives has not been reported in the literature to our knowledge.

Our study aimed to demonstrate whether additives consistent with food labels result in a significant amount of phosphorus in ready-to-eat flour tortillas such that their total phosphorus content would exceed that in ready-to-eat corn tortillas. Additionally, leavening agents comprising aluminum are commonly used to make ready-to-eat wheat but not ready-to-eat corn tortillas. Consistent with the ingredients on the label, aluminum additives are associated with high aluminum content in ready-to-eat flour tortillas. This is of concern in the general population and in CKD patients given the potential role of aluminum in neurological and neurodegenerative diseases including dialysis dementia.^20,21^

## Methods

Twenty-five different brands of tortillas, including 13 flour and 12 corn, were purchased in October 2019 and March 2020 from local supermarkets in Southern California including Target, food4less, Walmart, and Ralph’s, as well as fast food chains including Del Taco and Taco Bell. Each tortilla was assigned a code indicating whether it was corn- or flour-based and hard or soft and was weighed. The weights (mean ± SD) of the soft corn, soft wheat, and hard corn tortillas were 27.7 ± 7.7, 42.0 ± 12.6, and 15.5 ± 1.4 grams, respectively.

A sample weighing approximately 1 gram was removed from each tortilla (1.14 ± 0.09 g, mean ± SD). A subsample weighing about 250 mg was removed, dried to a constant mass, accurately weighed, and digested using trace-metal grade HNO_3_ in sealed Teflon vessels in a CEM MARS Express microwave digestion system (U.S. EPA method 6020), and analyzed by ICP-MS to quantify aluminum (Agilent 7900, Agilent Technologies, Inc., Santa Clarita, CA; U.S. EPA method 6020b) with germanium added as an internal standard and analyzed compared to NIST-traceable standards. The samples were also analyzed by ICP-OES (Agilent 5110 SVDV; U.S. EPA Method 6010d) to quantify phosphorus with yttrium added as an internal standard and analyzed compared to NIST-traceable standards. NIST standard reference material 1515, Apple Leaves, was used to verify the accuracy of the methods. The method detection limits (MDL) are shown in Table S1.

Each tortilla was tested for 21 elements (Ag, Al, As, Ca, Cd, Co, Cr, Cu, Fe, K, Mg, Mn, Na, Ni, P, Pb, Se, Sr, U, V, and Zn) (Table S1). Food labels, either on the packaging or from online searches, were compiled to identify the ingredients, focusing on food additives comprising phosphorus and aluminum. Multiple group comparisons were analyzed by running one-way ANOVA with Tukey’s post hoc test. Outliers were identified using the ROUT test on GraphPad/PRISM and statistical significance was determined using nonparametric Mann-Whitney and Kruskall-Wallis tests. All conditions (soft corn, hard corn, and wheat) were compared using Dunn’s multiple comparison test (Table S1).

### Justification for Sample Size

There are limited commercial suppliers of ready-to-eat tortillas, thereby limiting the sample size of this study. An effect size > 0.5, indicating a large difference effect seemed appropriate for this study.^22^ Most samples were taken in duplicate from the same tortilla package. Some samples were tested from a single source due to limited availability including a single homemade tortilla received as a gift. Analysis was standardized to grams per 30 gram serving since the size of each tortilla varied by brand. Statistical power was calculated using WebPower, statistical power analysis online.^23^

## Results

The elemental content of store-bought, ready-to-eat, wheat and corn tortillas was quantified. The elemental concentrations measured in this study were comparable to the calcium, iron, and potassium concentrations on the package label (Table S2). Further analysis of the phosphorus concentrations reveals that wheat flour tortillas generally contained approximately 50% more phosphorus (per 30 grams serving) than corn tortillas when a source of phosphorus was listed on the ingredients label (p=0.049). An effect size of 0.74 for the phosphorus content of wheat and soft corn tortillas with phosphorus additives indicates a practical significance between the two (Figure 1). Notably, wheat flour tortillas from the brands Old El Paso, La Banderita, Great Value, and Calidad contained approximately double the average phosphorus content of corn tortillas even when the latter listed phosphorus containing additives on the label (Table S3).

**Figure 1.**
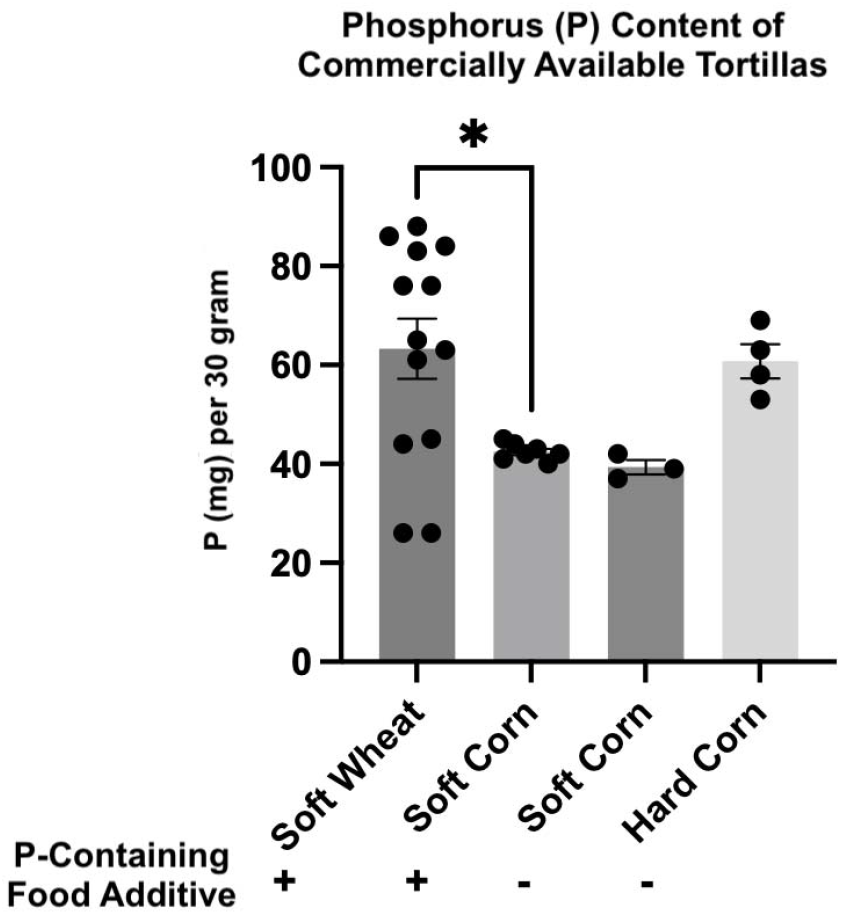
Total elemental phosphorus (P) content (mg P/30 grams serving) of commercially available, ready-to-eat tortillas, with or without P-containing food additives as per the label, including soft wheat, soft corn, and hard corn tortillas. *P<0.05; ANOVA p-value = 0.026; d = 0.89. Values are mean ± SEM.

The tested brands of corn tortillas had low aluminum content consistent with lack of aluminum-containing food additives reported on the label (Figure 2). On the other hand, wheat flour tortillas with reported aluminum additives had markedly elevated amounts of aluminum per 30 gram serving (p=0.0032 and p=0.0015 compared to wheat flour and corn tortillas with no added aluminum) (Figure 2). For wheat flour tortillas with aluminum additives compared to wheat without aluminum additives, soft corn, and hard corn tortillas, the effect sizes are 0.64, 0.69, and 0.56, suggesting that in wheat tortillas containing added aluminum the aluminum content is significantly higher compared to the other tortilla products. Surprisingly, many brands of wheat flour tortillas that did not list aluminum additives on the label also had an aluminum content comparable to Calidad brand tortillas that listed aluminum on the label (Table S4). In general, wheat flour tortillas contain more aluminum than corn tortillas even in the absence of aluminum additives listed on the label (0.46 ± 0.18 and 0.14 ± 0.11, respectively; mean ±SD) (Table S5).

**Figure 2.**
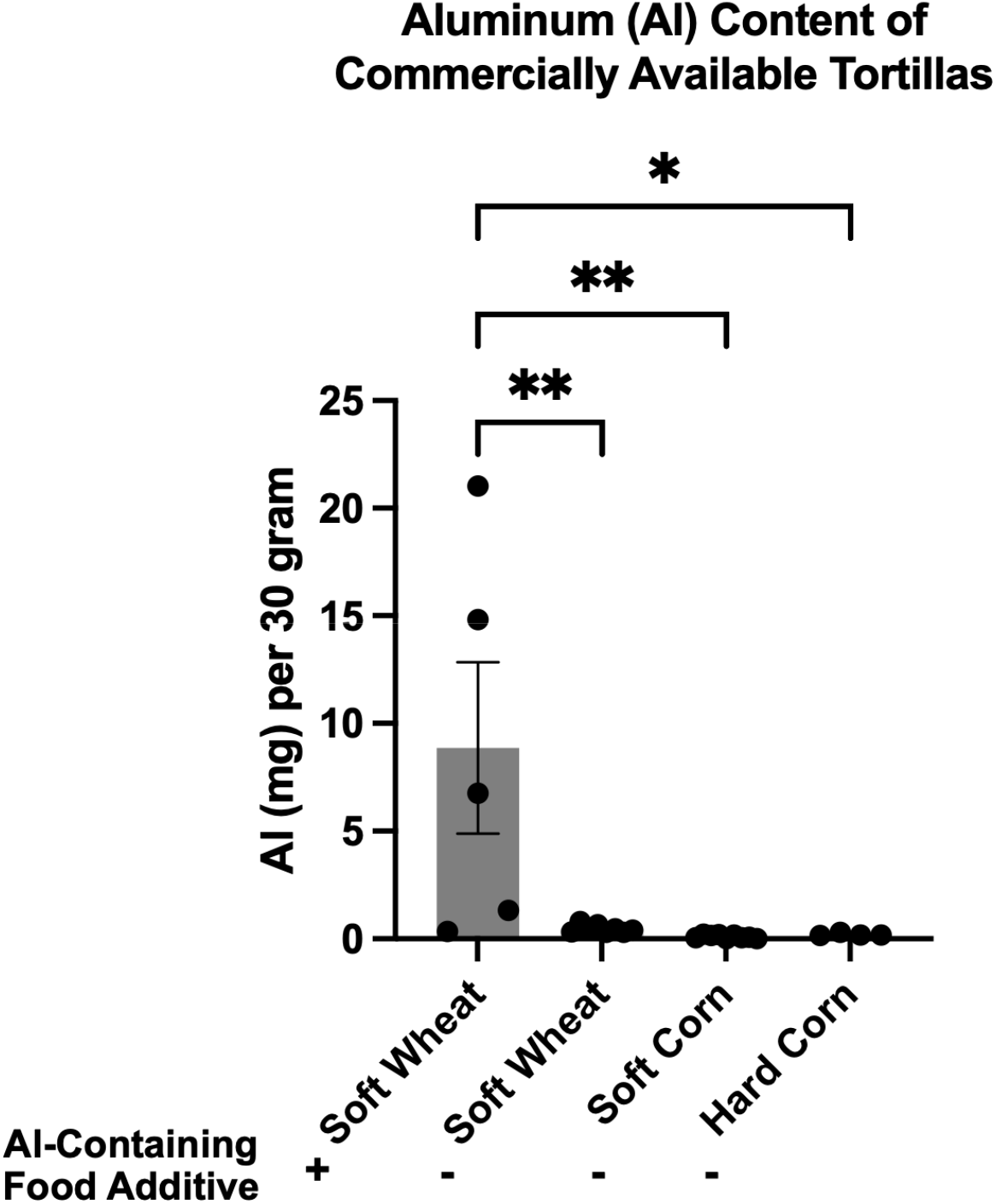
Aluminum (Al) content (mg Al/30 g serving) of commercially available, ready-to-eat tortillas, with or without Al-containing food additives as listed on the label, including soft wheat, soft corn, and hard corn tortillas. *P<0.05 **P<0.01; ANOVA p-value = 0.0013; d = 0.75. Wheat flour values are mean ± SEM.

Four brands of hard corn tortillas were studied: Del Taco, La Pericos, Old El Paso, and Taco Bell. The label on all four did not list any phosphorus- or aluminum-containing food additives. The phosphorus content ranged from 53 to 69 mg per 30 gram serving. This was comparable to wheat flour tortillas with phosphorus additives (63 ± 22 mg per 30 gram serving; mean ± SD) (Table S3). The aluminum content ranged from 0.15 to 0.31 mg per 30 gram serving, less than wheat flour tortillas with aluminum additives (8.86 ± 8.91 mg per 30 gram serving; mean ± S.D.) (Table S4) and comparable to soft corn tortillas (Table S5).

A single homemade wheat flour tortilla was tested. The phosphorus and aluminum concentrations were comparable to wheat flour tortillas that had added phosphorus but not aluminum salts (Table S1).

## Discussion

This study describes the aluminum and phosphorus content of ready-to-eat tortillas obtained from supermarkets and food franchises in Southern California and a homemade tortilla. The conventional wisdom has been that corn tortillas should be avoided in patients with chronic kidney disease because of the intrinsically high phosphorus content of corn.^24^ The website for Davita Kidney Care reports that the phosphorus content for a 30 gram portion for a six-inch corn tortilla is 75 mg while a flour tortilla made without baking powder is 20-37 mg.^25^ However, our findings show that wheat flour tortillas with approved food additives can have a phosphorus and aluminum content of up to about 90 mg and 20 mg per 30 gram serving, respectively. When elemental content is analyzed per tortilla, wheat flour tortillas consistently have significantly higher levels of phosphorus and aluminum compared to corn tortillas (p=0.005 and <0.0001, respectively) (Table S1). These findings are of great public health importance especially for countries with a large Hispanic population.

Homemade tortillas have a shelf-life of approximately 2-3 days while packaged flour and corn tortillas have a shelf-life of approximately 7 days after the expiration date.^26,27^ Although storage of tortillas at 4 °C is sufficient to prolong shelf life, limited refrigerator space has favored use of preservatives for storage at room temperature.^28^ The following preservatives are commonly added to tortillas: calcium propionate, propionic acid, sorbic acid, potassium sorbate, fumaric acid, citric acid, benzoic acid, phosphoric acid, lime, and sodium hydroxide. The addition of numerous preservatives, including phosphoric acid, sodium aluminum phosphate, sodium acid pyrophosphate, and calcium propionate, to flour tortillas may have contributed to higher levels of phosphorus in flour tortillas than in corn tortillas.

The daily dietary intake of an adult in the US comprises about 5 mg aluminum.^29,30^ One 30 gram serving of a wheat flour tortilla with aluminum reported on the label contains approximately 8.9 mg of aluminum. Given the per capita tortilla consumption of about 230-330 grams per day as reported from Mexico,^13^ the daily aluminum exposure would be about 68-98 mg of aluminum.

With a projected annual growth rate of 3.4%, the global commercial tortilla market may grow from 26 billion in 2022 to approximately 37 billion in 2032,^31^ thereby potentially increasing aluminum exposure and posing a serious health risk to patients with chronic kidney disease.

Sodium bicarbonate (baking soda), monocalcium phosphate, and either sodium acid pyrophosphate or sodium aluminum sulfate are often used as additives or preservatives in commercially-available tortillas.^32^ Baking powder is commonly used as a leavening agent in making bread and can be a source of aluminum.^33^ Even though not all tested brands listed aluminum on the label, we found that the range of aluminum content of wheat flour tortillas without added aluminum (0.31-14.83) overlaps with those that have added aluminum (0.31-21.04). It is possible that in products without listed aluminum on the label, aluminum was not deliberately added but rather was present in one of the additives. Water used to prepare tortillas may contain aluminum concentrations as high as 0.2 mg/L,^34^ but this would not be a significant additional source of aluminum. We also found a higher average aluminum content in wheat flour tortillas than in corn tortillas. Wheat flour tortillas contained about 0.030-0.330 mg aluminum per 30 gram serving depending on its source.^35-37^ Wheat flour has been reported to have a higher aluminum concentration compared to corn flour.^35,38,39^ Furthermore, wheat flour tortillas contained markedly higher levels of sodium compared to corn tortillas (Supplemental Table S1), likely due to sodium-based leavening agents.

The cereal and tuber food groups mainly comprising corn and flour tortillas respectively make up approximately 40% of the total energy intake of low-income Mexican agricultural workers.^40^

The average weight of the tortillas in this study was 35 grams. Given an average daily consumption of 10 and 7 tortillas by Hispanic males and females, respectively,^41-44^ consumption of some soft flour tortillas, e.g., Del Taco, Old El Paso, Romero’s, could easily exceed the TWI of 1.0 mg Al/kg body weight.^30^ High rates of chronic kidney disease of unknown origin have been frequently reported in agricultural workers known as mesoamerican nephropathy.^12^ Interestingly, toenail aluminum concentration was almost twice as high in those with acute Mesoamerican nephropathy than controls.^45^ In Central American workers with Mesoamerican nephropathy, increased consumption of corn and flour tortillas, especially store-bought ready-to-eat tortillas, raises the specter of aluminum toxicity and phosphorus associated cardiovascular disease.

Limitations of the current study include a small sample size that does not account for the diversity of tortilla brands globally and the different local preparation techniques for homemade tortillas. Further studies are needed to assess the aluminum and phosphorus content of homemade tortillas. Moreover, directly measuring the impact of tortilla intake on the aluminum and phosphorus levels in patients with kidney and cardiovascular disease would further guide dietary recommendations. The elemental concentrations measured in the current study were overall consistent with the available calcium, iron, potassium, and sodium levels reported on the packaging and ingredient labeling (Table S2). Our results indicate that patients and healthcare providers should be cautious of food labeling and limit intake of tortillas containing phosphorus and aluminum-based additives, particularly wheat flour tortillas.

## Conclusion

Contrary to conventional teaching, ready-to-eat flour tortillas have a higher elemental phosphorus concentration than corn (maize) tortillas due to added preservatives and leavening agents. Use of aluminum based leavening agents to make ready-to-eat wheat tortillas confers a higher aluminum concentration compared to corn tortillas. The aluminum and phosphorus concentrations of many ready-to-eat tortilla brands can present a daily dietary load of up to approximately 100 mg aluminum and 700 mg phosphorus based on an average daily tortilla intake of 330 grams. Despite conventional wisdom that CKD patients should avoid phosphorus rich corn tortillas, ready-to-eat wheat flour tortillas consistently had a higher aluminum and phosphorus content due to additives. Patients with CKD should favor homemade tortillas rather than ready-to-eat tortillas and pay close attention to additives listed on the label.

## Data Availability

All data produced in the present study are available upon reasonable request to the authors.

## Abbreviations

P: Phosphorus
Al: aluminum
CKD: chronic kidney disease
TWI: tolerable weekly intake
ICP-MS: inductively coupled plasma mass spectrometry
ICP-OES: inductively coupled plasma optical emission spectroscopy

## Acknowledgements

The authors gratefully acknowledge Marsha L. Ensor and Shristi Shrestha for their contributions to this research.

This publication was supported by UK-CARES through Grant P30ES026529. Its contents are solely the responsibility of the authors and do not necessarily represent the official views of the NIEHS.

## Author Contributions

A.G., K.C.C. and R.Y. created the framework for the manuscript for the manuscript and wrote the original draft. JU conducted sample testing and wrote the methods for the manuscript. All authors have reviewed and agreed on the final version of the manuscript.

