## Supplemental Tables 3-5 for "Dietary burden of phosphorus and aluminum in ready-to-eat wheat flour tortillas exceed that of corn tortillas: Implications for patients with renal or cardiovascular disease"

**Table S1.** The maize and wheat/flour tortillas studied, their sources, hard or soft composition, total weight, and elemental content (See Supplemental File 1)

**Table S2.** Elemental concentrations measured per tortillas compared to listed concentrations (See Supplemental File 2)

**Table S3: Phosphorus (P) content of wheat flour and corn tortillas with phosphorus-containing food additives as per the label**.

|  | **Brand** | **P-containing food additives** | **P (mg) per 30 g serving** |
| --- | --- | --- | --- |
| **Wheat Flour Tortillas** |  |  |  |
|  | Calidad | Na_2_H_2_P_2_O_7_, Ca(H_2_PO_4_)_2_ | 83 |
|  | Diana’s | Al_3_H_22_NaO_36_P_8_ | 26 |
|  | Del Taco | Na_2_H_2_P_2_O_7_, Ca(H_2_PO_4_)_2_ | 26 |
|  | El Comal | Na_2_H_2_P_2_O_7_, Ca(H_2_PO_4_)_2_ | 63 |
|  | Guerrero | Na_2_H_2_P_2_O_7_ | 44 |
|  | Great Value | Na_2_H_2_P_2_O_7_ | 86 |
|  | Kroger | Na_2_H_2_P_2_O_7_ | 76 |
|  | La Banderita | Na_2_H_2_P_2_O_7_, Ca(H_2_PO_4_)_2_ | 84 |
|  | Mission | Na_2_H_2_P_2_O_7_ | 65 |
|  | Más y Más | Na_2_H_2_P_2_O_7_ | 76 |
|  | Old El Paso | Al_3_H_22_NaO_36_P_8_ | 88 |
|  | Romero’s | Na_2_H_2_P_2_O_7_ | 45 |
|  | Taco Bell | Na_2_H_2_P_2_O_7_, CaHPO_4_ | 61 |
|  | **Mean (SD)** | 63 (22) | |
| **Corn Tortillas** | Calidad | H_3_PO_4_ | 42 |
|  | Del Taco | H_3_PO_4_ | 45 |
|  | El Comal | H_3_PO_4_ | 40 |
|  | Guerrero | H_3_PO_4_ | 43 |
|  | Great Value | H_3_PO_4_ | 44 |
|  | Mission | H_3_PO_4_ | 41 |
|  | Romero’s | H_3_PO_4_ | 42 |
|  | **Mean (SD)** | 42 (2) | |
| **Flour vs Corn Tortillas** | **Ratio (flour/corn)** | 1.49 | |
|  | **% increase*** | 50% | |

*[(P in flour-P in corn)/P in corn] x 100

P refers to inorganic phosphorus, including P in the grain. Total P was determined analytically.

**Table S4: Aluminum content of wheat flour tortillas with or without aluminum-containing food additives as per the label**

|  | **Brand** | **Al-containing food additives** | **Al (mg) per 30 g serving^** |
| --- | --- | --- | --- |
| **Al-containing food additives added as per label** | Calidad | NaAl(SO_4_)_2_ | 0.35 |
|  | Diana’s | NaAl(SO_4_)_2_ Al_3_H_22_NaO_36_P_8_ | 1.33 |
|  | Del Taco | NaAl(SO_4_)_2_ | 14.83 |
|  | Old El Paso | Al_3_H_22_NaO_36_P_8_ | 21.04 |
|  | Romero’s | NaAl(SO_4_)_2_ | 6.77 |
|  | **Mean (SD)** | 8.86 (8.91) | |
| **Aluminum Ingredient-free as per label** | El Comal | - | 0.65 |
|  | Guerrero | - | 0.31 |
|  | Great Value | - | 0.47 |
|  | Kroger | - | 0.41 |
|  | La Banderita | - | 0.31 |
|  | Mission | - | 0.33 |
|  | Más y Más | - | 0.43 |
|  | Taco Bell | - | 0.80 |
|  | **Mean (SD)** | 0.46 (0.18) | |
| **Al added vs Al not added^**^** | **Ratio (Al added/Al not added)** | ~19 | |
|  | **% increase^+^** | 1800% | |

^This is the average value of duplicate measurements on each sample

**As per the ingredients listed by the manufacturer in the label

^+^[(Al added-Al not added)/Al not added] x 100

**Table S5: Aluminum content of tortillas when aluminum-containing food additives are not listed as ingredients on the label**

| **Brand** | | **Al (mg/30 g serving)** |
| --- | --- | --- |
| **Flour Tortillas** | | |
| El Comal | | 0.65 |
| Guerrero | | 0.31 |
| Great Value | | 0.47 |
| Kroger | | 0.41 |
| La Banderita | | 0.31 |
| Mission | | 0.33 |
| Más y Más | | 0.43 |
| Taco Bell | | 0.80 |
| **Mean (SD)** | | 0.46 (0.18) |
| **Corn Tortillas** | | |
| **Soft** | Calidad | 0.21 |
|  | Del Taco | 0.15 |
|  | El Comal | 0.17 |
|  | El Milagro | 0.08 |
|  | Guerrero | 0.02 |
|  | Great Value | 0.06 |
|  | La Banderita | 0.02 |
|  | Mission | 0.19 |
|  | Romero's | 0.06 |
| **Hard** | Del Taco | 0.15 |
|  | La Pericos | 0.17 |
|  | Old El Paso | 0.17 |
|  | Taco Bell | 0.31 |
| **Mean (SD)** | | 0.14 (0.11) |
| **Flour vs Corn Tortillas** | | |
| **Ratio (Al in flour/Al in corn)** | | 3.407 |
| **% Increase** | | 240% |

*****[(Al in flour-Al in corn)/Al in corn] x 100
